# Review of evidence regarding attributes and behaviours of smokers as smoking prevalence falls, including evidence relevant to the ‘hardening hypothesis’

**DOI:** 10.1101/2020.09.16.20195560

**Authors:** Miranda Harris, Melonie Martin, Amelia Yazidjoglou, Laura Ford, Robyn M Lucas, Emily Banks

## Abstract

To review evidence relevant to Australia and similar high-income countries regarding continuing smokers’ motivation, dependence and quitting behaviour as smoking prevalence declines, to assess whether population “hardening” (decreasing propensity to quit) or “softening” (the converse) is occurring. MEDLINE, PsychINFO, Scopus, Web of Science and Cochrane Library were searched to July 2019, using terms related to smoking and hardening, for reviews and large, population-based repeat cross-sectional studies. There were additional searches of reference lists and citations of key research articles. Two reviewers screened half the titles and abstracts each, and two reviewers screened all full texts independently using tested criteria. Four reviewers independently and systematically extracted data from eligible publications, with one reviewer per study, checked by another reviewer. Of 265 titles identified, three reviews and ten repeat cross-sectional studies (not included in the reviews) were included. All three reviews concluded that hardening has not occurred among the general smoking population over time. Of the ten repeated cross-sectional studies, five examined motivation, nine examined dependence, five examined hardcore smoking, and two examined quit outcomes over time. All found a lack of hardening and most found softening within the smoking population, consistent across hardening indicators, definitions, countries (and tobacco control environments) and time periods examined. Declining smoking prevalence has been accompanied by softening within the population of smokers, characterised by increasing motivation to quit and reduced dependency. Based on the weight of the available evidence from high-income countries, the “hardening hypothesis” should be rejected.

## Introduction

Tobacco use is the leading cause of preventable death and disability in Australia. The Australian Government has committed to reducing smoking prevalence to below 10% by 2025.^1^ The “hardening hypothesis” proposes that as the prevalence of smoking in a population declines, there will be a “hardening”, whereby smokers who are more resistant to cessation make up a greater proportion of the remaining smoker population.^2 3^ The hypothesis is based on the expressed concern that pressures to quit smoking from tobacco control policies and increasing social stigma of smoking could mean that smokers who found it relatively easy to quit would most readily cease smoking, and the smokers left behind would be increasingly resistant to tobacco control measures.^4^ The term “softening” has been coined to describe the opposite of hardening, whereby the smoking population displays behaviours characteristic of increasing willingness and/or ability to quit over time.

Indicators of hardening or softening can be categorised as measuring motivational or dependence hardening, proportion of hard-core smokers, and quit outcomes (Table 1).^5^ While socioeconomic disadvantage and psychological distress among smokers have been postulated to be indicators of hardening, these are not direct measures of hardening.^6^

**Table 1:** Constructs and measures of hardening among current smokers.

| **Hardening Constructs** | **Indicators** |
| --- | --- |
| Motivational hardening | Attitudes to smoking or tobacco control<br>Quit attempts<br>Quit intentions |
| Dependence hardening | Proportion of smokers who are daily smokers<br>Proportion of smokers who are heavy smokers<br>Mean cigarettes per day, among smokers<br>Questionnaire measures of dependence (e.g. Nicotine Dependence Syndrome Scale) |
| Hard-core smoker | A composite of the indicators of motivational and dependence hardening above |
| Quit outcomes | Success on a given quit attempt or ability to remain abstinent on a given quit attempt<br>Quit ratio (ratio of former smokers to ever smokers in a given population)<br>Proportion of the eligible smoking population who have quit in a given time period |

It should be noted that the hardening hypothesis is not a confirmed phenomenon and many tobacco control policies positively affect measures that are included in the concept of “hardening”. For example, increasing costs of tobacco products, restrictions on places where people can smoke, graphic health warnings and media campaigns all affect motivation and, along with support for cessation and reduced overall community prevalence of smoking, can improve quit outcomes. Reduction in the ability to smoke large numbers of cigarettes per day – including due to cost, restrictions on places where smoking is allowed and lack of social acceptability – is likely to affect dependence.

One hypothetical concern is that previously effective population-level interventions could become less successful if a population hardens, requiring greater emphasis on individual-level cessation interventions to reach hardened smokers who would make up a greater proportion of the population of smokers.^3 5 7^ Evidence of hardening would also be considered to provide potential justification for long-term nicotine replacement approaches – such as patches, gums and e-cigarettes – as “harm reduction” for smokers who struggle to quit nicotine use.^2 4 8^

The hardening hypothesis is most rigorously tested by examining changes in hardening or softening indicators within the population of smokers over extended periods of time, using a cohort or repeat cross-sectional study design.^5^ This review aims to summarise the contemporary evidence assessing the evidence for hardening or softening within the population of smokers in Australia and other high-income countries.

## Methods

### Definitions of hardening constructs and indicators

Motivational hardening may occur if the population of smokers become, on average, less motivated or willing to quit.^3 5^ Less motivated smokers are characterised by the absence of quit attempts or the lack of an intention to quit.^3^ A smoker’s attitude towards tobacco control measures has been proposed as an indirect measure of motivation to quit.^5^

Dependence hardening occurs if an increasing proportion of smokers are dependent (either physiologically dependent on nicotine or behaviourally on smoking).^3^ These smokers may experience multiple failed quit attempts and/or exhibit behaviour consistent with high levels of dependence such as heavy consumption, smoking soon after waking (measured by time to first cigarette), and high scores on questionnaires measuring dependence.^3^ The average number of cigarettes smoked per day has been used to measure whether the average dependence of smokers is changing. Multiple unsuccessful quit attempts is also considered a marker of dependence.^5 9^

A hard-core smoker is usually conceptualised as a smoker who is highly unwilling and/or unable to quit and likely to remain this way.^3^ Although there is no agreed definition of a hard-core smoker, the categorisation generally relates to both very low levels of motivation and very high levels of dependence.^10^ Common indicators used include nicotine dependence, regular smoking, lack of motivation or readiness to quit, and lack of recent quit attempts.^10^ Most definitions exclude smokers 25 years and younger, as these individuals are still establishing their smoking patterns.^10-12^ The concept of a hard-core smoker is an individual measure and is separate, but often related, to hardening, which is a population measure. It is possible to have hard-core smokers in a smoking population that does not show evidence of hardening over time. Conversely, the population of smokers may be hardening over time but the proportion of hard-core smokers may not change. These concepts are often linked in the published evidence in that the proportion of smokers who are classed as “hard-core” has been considered an indicator of hardening or softening of the smoking population.^3^

If hardening of the population of smokers were occurring due to reduced motivation or increased dependence, there would be a decline over time in the conversion of current smokers to former smokers.^5^ This is often measured by the “quit ratio” – the ratio of former smokers to ever smokers in a given population – or by the proportion of the eligible smoking population who have quit within the last twelve months.^3^ Success on a given quit attempt could also be considered a quit outcome.

### Literature search, screening and data extraction

Reviews and primary research studies of repeat large population-based cross-sectional studies from Australia, and countries similar to Australia, with a gap of at least 5 years between data points, in line with another review of hardening,^9^ were identified through a combination of database searches and reference and citation searches. MEDLINE, PsychINFO, Scopus, Web of Science and Cochrane Library were searched up until July 2019 using a search strategy detailed in Supplementary material 1.

Two review authors screened half the titles and abstracts each, independently. Two review authors screened all full texts using tested criteria, with disagreement about eligibility resolved through discussion involving a third reviewer. Studies were excluded if they were not representative of the general population or had less than 1,000 participants for any survey year (full inclusion/exclusion criteria in Supplementary material 2).

Four review authors independently extracted data from studies using piloted data extraction spreadsheets, with a check performed by another reviewer. The quality of included repeated cross-sectional studies was independently assessed by four review authors (two per study) using a tool adapted from the Joanna Briggs Institute Critical Appraisal Checklist for Studies Reporting Prevalence Data and the National Heart, Lung, and Blood Institute Study Quality Assessment Tool for Observational Cohort and Cross-Sectional Studies.^13 14^ As no systematic reviews were identified, the quality of the included reviews was not assessed. Disagreements were resolved through discussion between the two review authors, and through discussion involving a third reviewer when required. Author declarations of interest and other relevant information were reviewed and summarised. For interpretability, where relevant, this review reports on the change in the proportion of smokers meeting the hardening indicator definition over time.

## Results

Of 265 titles identified, three reviews and ten repeat cross-sectional studies were identified for inclusion (Figure 1).

**Figure 1:**
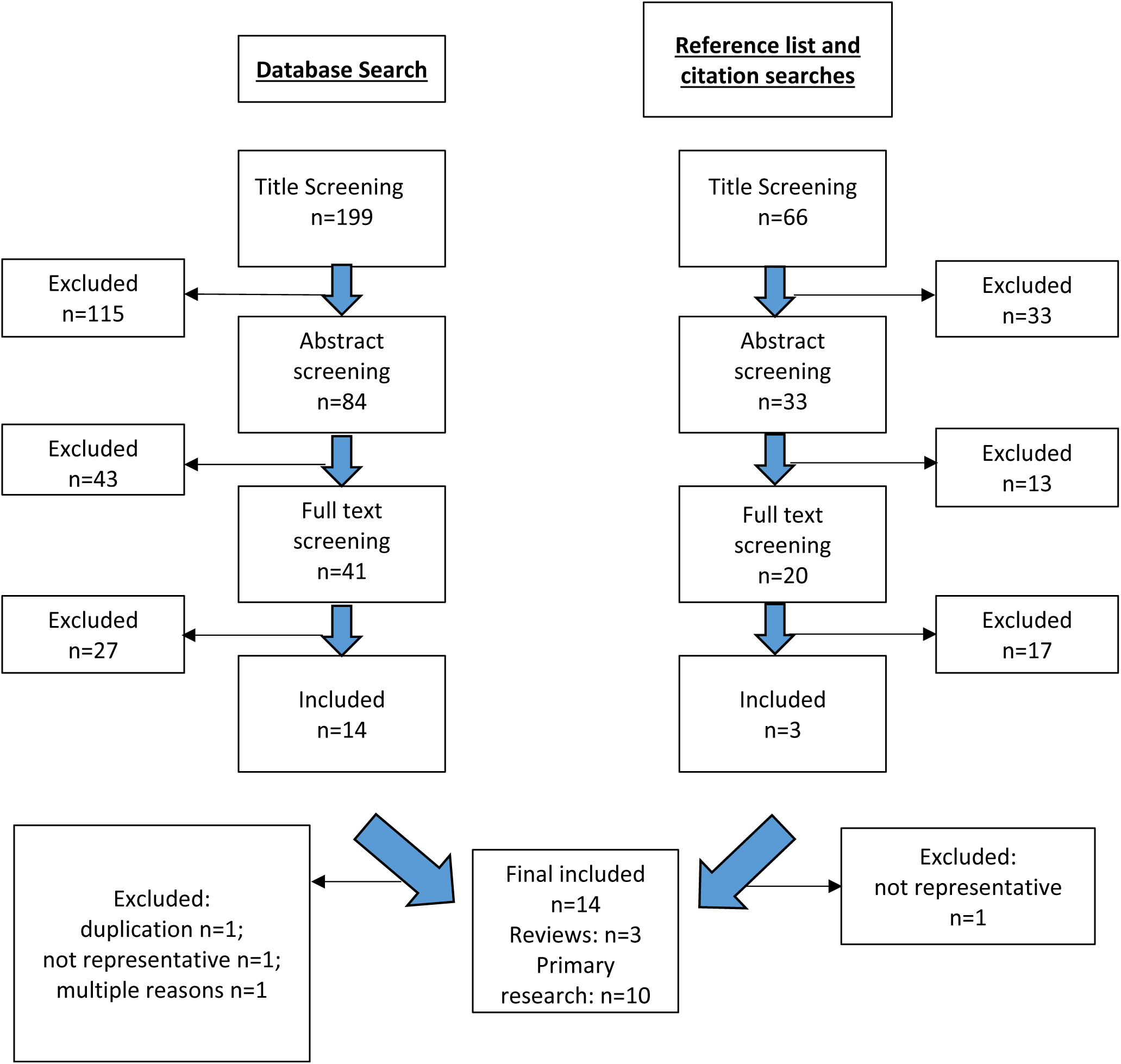
Flow diagram demonstrating study selection.

### Reviews

All three reviews conclude that hardening has not occurred among the general population of smokers, despite each considering different evidence.^3 6 9^ In 2003, Warner and Burns^3^ reviewed three empirical analyses on hardening and presented evidence against the case of hardening in the US population. They concluded that the proportion of hard-core smokers in the population was very small and at the time of their review, there was little evidence that hardening was occurring at the population level.^3^ In 2011, Hughes^6^ updated the review by Warner and Burns^3^ and a review from the US Department of Health and Human Services,^15^ identifying two new studies on quit attempts, plotting quit ratios from the US National Health Interview Supplement, and reviewing five new studies relating to nicotine dependence. Hughes^6^ found no evidence of hardening among the general population of smokers, but did find evidence of hardening among treatment seekers. In 2019, Hughes^9^ undertook another review of 26 studies to assess whether there was a decrease over time in (1) conversion from current to former smoking; (2) quit attempts; or (3) success on a given quit attempt. None of the reviewed studies found evidence of hardening, and many found evidence of softening (Supplementary material 3). Hughes (2019)^9^ provides the most recent and robust review of hardening, but does not include data on quit intentions, dependence, and attitudes on tobacco control.

### Primary evidence

None of the ten repeated cross-sectional studies included had been considered in the reviews by Warner and Burns,^3^ Hughes (2011),^6^ or Hughes (2019).^9^ Although one of the studies included in Hughes (2019)^9^ has the same reference as Kulik and Glantz (2016)^16^ which is included in the current review, based on the information presented in Hughes (2019), it does not appear to be the same study. One study,^17^ which was excluded from the review of primary research, set out to replicate and critique the findings of another included study.^16^ Reasons for excluding this study were: duplication of data from the original study; adjustment for variables such as tobacco control policy that were highly correlated with smoking prevalence and likely to be mediators of softening over time; and potential competing interests of the authors.^17 18^

Of the ten repeated cross-sectional studies included, five examined motivation, nine examined measures of dependence, five examined hard-core smoking, and two examined quit outcomes over time. Eight studies examined hardening in the 2000s,^5 11 19-24^ and two examined hardening across both the 1990s and the 2000s. Four studies were conducted in the US,^16 22-24^ with one of these studies also examining 31 countries across Europe.^16^ The latter study^16^ presented analyses of European data for two hardening indicators; only one of these analyses has been reported in this review as the other analysis did not meet the minimum time period between data points required for inclusion. Two studies were conducted in Australia^20 21^ and there was one study each from New Zealand,^5^ Canada,^19^ Norway^25^ and England.^11^

An overview summary of the results is presented in Table 2, with detailed findings presented in Supplementary material 4. Supplementary material 5 provides a summary of authors’ conflicts of interests.

**Table 2:** Overview of the results from 10 repeated cross-sectional studies in Australia and similar countries examining hardening indicators

| Authors | Indicator | Finding |
| --- | --- | --- |
| **Motivation – Quit intention** |  |  |
| **Australia** |  |  |
| Clare et al (2014)<sup>21</sup> | No plan to quit | Softening |
| Brennan et al (2019)<sup>20</sup> | No intention to quit in the next 30 days | Softening |
| Brennan et al (2019)<sup>20</sup> | No intention to quit in the next 6 months | Softening |
| Brennan et al (2019)<sup>20</sup> | Happy to smoke for the rest of their lives | Softening |
| **International** |  |  |
| Docherty et al (2014)<sup>11</sup> | Low motivation to quit | Neither hardening nor softening |
| **Motivation – Quit attempts** |  |  |
| **Australia** |  |  |
| Clare et al (2014)<sup>21</sup> | No quit attempt in the past 12 months | No statistical test reported (the authors reported that the proportion of smokers with no quit attempt in the past 12 months was consistent across the four waves) |
| Brennan et al (2019)<sup>20</sup> | No quit attempt in the past 12 months | Softening |
| Brennan et al (2019)<sup>20</sup> | No quit attempt in the past 5 years | Softening |
| Brennan et al (2019)<sup>20</sup> | Never attempted to quit | Neither hardening nor softening |
| **International** |  |  |
| Kulik and Glantz (2016)<sup>16</sup> | Made a quit attempt in the past 12 months (US) | Softening |
| Kulik and Glantz (2016)<sup>16</sup> | Made a quit attempt in the past 12 months (EU) | Neither hardening nor softening |
| Edwards et al (2017)<sup>5</sup> | No quit attempts in the past 12 months | Neither hardening nor softening |
| **Motivation – Attitudes to tobacco control** |  |  |
| **International** |  |  |
| Edwards et al (2017)<sup>5</sup> | Agree with banning smoking in all public places where children are likely to go | Softening |
| Edwards et al (2017)<sup>5</sup> | Agree the number of places allowed to sell cigarettes and tobacco should be reduced | Neither hardening nor softening |
| Edwards et al (2017)<sup>5</sup> | Supported cigarettes and tobacco should not be sold in New Zealand in 10 years' time | Neither hardening nor softening |
| **Dependence** |  |  |
| **Australia** |  |  |
| Clare et al (2014)<sup>21</sup> | Heavy smoking | No statistical test reported (the authors reported that the proportion of heavy smokers was consistent across the four waves) |
| Brennan et al (2019)<sup>20</sup> | Daily smoking | Softening |
| Brennan et al (2019)<sup>20</sup> | Heavy smoking | Softening |
| **International** |  |  |
| Coady et al (2012)<sup>22</sup> | Cigarettes per day in current smokers | Softening |
| Coady et al (2012)<sup>22</sup> | Heavy daily smoking | Softening |
| Docherty et al (2014)<sup>11</sup> | Time to first cigarette ≤ 30 minutes after waking | Neither hardening nor softening |
| Docherty et al (2014)<sup>11</sup> | Time to first cigarette ≥ 30 minutes after waking | Neither hardening nor softening |
| Smith et al (2014)<sup>24</sup> | Heavy smoking | No statistical test reported |
| Smith et al (2014)<sup>24</sup> | Nicotine Dependence Syndrome Scale scores | Softening |
| Azagba (2015)<sup>19</sup> | Time to first cigarette ≤ 5 minutes and/or ≤ 30 minutes after waking | Neither hardening nor softening |
| Kulik and Glantz (2016)<sup>16</sup> | Cigarettes per day in current smokers | Softening |
| Edwards et al (2017)<sup>5</sup> | Daily smoking | Neither hardening nor softening |
| Edwards et al (2017)<sup>5</sup> | Daily smoking with 4 or more quit attempts in the past 12 months | Neither hardening nor softening |
| Goodwin et al (2018)<sup>23</sup> | Heavy smoking | Softening |
| Goodwin et al (2018)<sup>23</sup> | Time to first cigarette < 30 minutes after waking | Softening |
| **Hard-core smokers** |  |  |
| **Australia** |  |  |
| Clare et al (2014)<sup>21</sup> | Hard-core smokers | Neither hardening nor softening |
| Brennan et al (2019)<sup>20</sup> | Hard-core smokers | Softening |
| Brennan et al (2019)<sup>20</sup> | Given up giving up | No statistical test conducted |
| **International** |  |  |
| Lund et al (2011)<sup>25</sup> | Daily heavy smoking | Softening |
| Docherty et al (2014)<sup>11</sup> | Hard-core smokers | Hardening |
| Azagba (2015)<sup>19</sup> | Hard-core smokers | Neither hardening nor softening |
| **Quit outcomes** |  |  |
| **International** |  |  |
| Kulik and Glantz (2016)<sup>16</sup> | Quit ratio | Softening |
| Edwards et al (2017)<sup>5</sup> | Recent quit rate | Neither hardening nor softening |
| Edwards et al (2017)<sup>5</sup> | Recent sustained quit rate | Neither hardening nor softening |

### Motivation

Five studies examined change in motivation over time, including the two Australian studies. Analysis of a series of large national household surveys in Australia shows the odds of having no plans to quit were significantly lower in 2010 compared with all previous years (odds ratio (OR) 0.87, 95% CI 0.77-0.98).^20^ The proportion of smokers in a subsequent state-based study in Victoria, Australia also shows that smokers were less likely over time to have no intention to quit, with those who had no intention of quitting in the next 30 days or the next six months decreasing between 2001 and 2016 (adjusted OR (aOR) per calendar year 0.95, 95% CI 0.93-0.96, p(trend) < 0.001, and aOR per calendar year 0.97, 95% CI 0.96-0.98, p(trend) < 0.001 respectively).^20^ The proportion of smokers attempting to quit either remained consistent or significantly declined.^20 21^ The Victorian study also found that there was a significant decrease in the proportion of smokers who indicated they were happy to smoke for the rest of their lives (aOR per calendar year 0.97, 95% CI 0.95-0.99, p(trend) = 0.001).^20^

International evidence on quit intentions and attempts suggests that as smoking prevalence declines, the smoking population is either becoming more motivated to quit, or remaining stable in its motivation.^5 11 16 17^ New Zealand smokers’ attitudes to tobacco control measures and goals, as a proxy measure for motivation, have softened over time or remained unchanged.^5^ Between 2008 and 2014, there was a steady increase over time in the proportion of daily smokers who supported banning smoking in all public places where children are likely to go (2008: 44.8%, 2014: 66.3%; aOR per two-year increment 1.16, 95% CI 1.08-1.25, p(trend) not reported).^5^ The proportion of daily smokers who agreed with reducing the number of places allowed to sell tobacco to make it less available showed no significant change, as did support for cigarettes and tobacco not being sold in New Zealand in ten years’ time.

### Dependence

Two studies in Australia,^20 21^ four in the US,^16 22-24^ and one in Canada,^19^ England^11^ and New Zealand^5^ examined change in markers of dependence over time. The measures used to examine dependence differed across studies, with cigarettes per day being the most common measure and the proportion of either heavy smokers or daily smokers within the smoking population also frequently used. The definition of heavy smoking varied; some publications defined heavy smoking as at least 15 or 16 cigarettes per day while another used at least 25 cigarettes per day as the threshold. Other measures included time to first cigarette after waking and the Nicotine Dependence Syndrome Scale.

In Australia, the Victorian study found that smokers were increasingly less likely to be daily or heavy smokers between 2001 and 2016 (84.2% to 79.7%, aOR per calendar year 0.96, 95% CI 0.95-0.98; p(trend) < 0.001),^20^ whilst the national study found no change between 2001 and 2010 (noting that no statistical test was reported for the dependence measure).^21^ Thresholds for heavy smoking were similar for the two studies: 15 cigarettes per day and 16 cigarettes per day, respectively. Neither of these Australian studies reported the prevalence of heavy smoking in the total population, however these values were calculated from their data. The estimated prevalence of heavy smoking in the total population was 9.4% in 2001 and 8.0% in 2010 in Australian adults aged 18 years and over, and 8.5% in 2001 and 2.8% in 2016 in Victorian adults aged 26 years and over.^20 21^ The Victorian study found no variation in the change in prevalence of heavy smoking over time according to age, sex, education or socioeconomic status.^20^

International studies suggest that dependence is on average declining or not changing in smokers, demonstrated by a decrease or no change in the proportion of smokers who were daily or heavy smokers,^5 22-24^ a decrease or no change in the proportion of smokers who were smoking soon after waking,^11 19 23^ no change in the proportion of smokers with four or more quit attempts of more than 24 hours in the past year^5^ and a decrease in dependence scores on the Nicotine Dependence Severity Scale.^24^ Of the smokers that continued to smoke, consumption, measured by average number of cigarettes per day, declined over time.^16 17^ In their US study, Smith et al^24^ examined sociodemographic factors and comorbidities, finding that declines in dependence severity (on the Nicotine Dependence Severity Scale) were greatest for smokers without any serious psychological distress. No significant variation in change in dependence severity over time was found according to sex, annual income or age.^24^

### Hard-core smoking

Five studies examined hardening in the population of smokers over time based on data related to hard-core smoking.^11 19-21 25^ In Australia, there was no evidence of hardening as measured by change in proportion of the smoking population who were hard-core smokers. The Victorian study found a significant decline in the proportion of smokers who were hard-core between 2001 and 2016 (17.2% to 9.1% aOR per calendar year 0.94, 95% CI 0.92-0.96; p(trend) < 0.001),^20^ and the national study found no significant change over four waves from 2001 to 2010 (2001: 11.9%, 2004: 10.9%, 2007: 11.8%, 2010: 10.7% p(heterogeneity by wave) = 0.550).^21^ The overall population prevalence of hard-core smoking was 2.5% in 2000 and 2.0% in 2010 using data from the Australian study by Clare et al^21^ (no statistical tests undertaken). The population prevalence of hard-core smoking was estimated as 3.2% in 2001 and 1.2% in 2016 in the adult Victorian population aged 26 years and older (no statistical tests undertaken).^20^ The hard-core smoker definitions used in these two studies were similar. Within the Victorian population, the proportion of the “given up giving up” group (defined as daily smokers who had previously made five or more quit attempts, had not made a quit attempt within the past five years, or who did not intend to quit within the next six months) was calculated to be around 0.2% of adult Victorians in 2001 and 0.1% in 2016.

Brennan et al^20^ undertook sensitivity analyses to explore the impact on the findings of using different definitions of hard-core smoker. In one definition, the criterion of not making a quit attempt within the past twelve months was replaced with having never attempted to quit. In two additional analyses, the heavy consumption criterion was removed as authors noted that cigarette consumption may be influenced by tobacco control policies, such as smoke-free policies, reducing the opportunities to smoke rather than reflecting the nicotine dependence of an individual. Regardless of the definition used, the proportion of smokers who were hard-core smokers decreased significantly over time, supporting the findings of the primary analysis.

Using national Australian data, Clare et al^21^ found that the change in the proportion of hard-core smokers over time varied according to socioeconomic status (p(interaction) = 0.025); between 2001 and 2010, the proportion of being a hard-core smoker declined among people of higher socioeconomic status (2001: 9.3%, 2010: 6.7%) but remained static among those of lower socioeconomic status (2001: 13.7%, 2010: 13.7%). Victorian data^20^ also indicate a difference in changes in the proportion of hard-core smokers over time by level of education (p(interaction) < 0.017); however this was not significant at the authors’ pre-specified p ≤ 0.01 level. The decline over time in the proportion of hard-core smokers (aOR per calendar year 0.97, 95% CI: 0.94-0.99, p(trend) = 0.012) was smaller in the group with lower education compared to that in the higher education group (aOR per calendar year 0.92, 95% CI: 0.90-0.94, p(trend) < 0.001). The proportion of hard-core smokers did not differ over time according to an area-based measure of socioeconomic status (p(interaction) = 0.434).

A Norwegian study using nationally representative data found evidence of softening, demonstrated by a decline in the proportion of smokers who were hard-core smokers over the period 1996 to 2009 (OR per increment in survey year (2 years) 0.90, 95% CI 0.88-0.93).^25^ There was no evidence of a change in the proportion of smokers who were hard-core in Canada between 2004 and 2010 using nationally representative data.^19^

An English study assessed data from two national datasets in England, both analyses finding there was an increase in the proportion of smokers who were defined as hard-core in England between 2000 and 2010 (UK General Lifestyle Survey p(trend) < 0.001; Health Survey for England p(trend) = 0.04).^11^ However, when the two components of the hard-core smoker definition were examined separately, there was no statistically significant change over time in either survey in the odds of smokers who did not want to quit (p(trend) = 0.760 and 0.592 respectively), or for smokers who had their first cigarette within 30 minutes after waking (p(trend) = 0.288 and 0.785 respectively). Based on graphs presented by the authors, the proportion of smokers who were hard-core was estimated to have increased by approximately 1% and 2% in the General Lifestyle Survey, and the Health Survey for England, respectively, over the eleven-year time period.

### Quitting outcomes

One US study^16^ included an examination of the relationship between quit ratio and smoking prevalence and one New Zealand Study^5^ examined recent and sustained quit rates. Quit outcomes were not examined in either of the Australian studies. The US study found that the quit ratio increased as smoking prevalence declined between 1992/93 and 2010/11: an increase of 1.13% (± 0.06 standard error, p < 0.001) for each 1% decrease in smoking prevalence.^16^ In the New Zealand study, authors found no significant change between 2008 and 2014 in recent quit rates (2008: 8.4%, 2014: 9.5%; aOR per two-year increment 1.03 (95% CI: 0.92-1.15)) or recent sustained quit rates (2008: 6.9%, 2014: 12.4%; aOR per two-year increment 1.12 (95% CI: 0.96-1.30)).^5^

## Discussion

There is no evidence of hardening of the smoking populations in the countries examined, including Australia, with virtually all indicators consistent with softening or showing no significant change. The available evidence indicates that between 2001 and 2016, the Australian population of smokers has become, on average, more motivated to quit and less dependent on smoking. In countries similar to Australia (studies from the US, Norway, England, Europe, New Zealand, and Canada), the available evidence does not indicate hardening of the population of smokers between 1992 and 2015, and in many cases shows softening. The findings are consistent with the reviews by Warner and Burns,^3^ Hughes (2011)^6^ and Hughes (2019),^9^ which did not find evidence of hardening. Based on the evidence to date, the lack of hardening within the population of smokers is almost completely consistent across the range of hardening indicators employed, their definitions, countries (and tobacco control environments) and time periods examined.

The hardening hypothesis and the concept of hard-core smoking are part of a broader way of thinking about tobacco control, and are increasingly being questioned, in view of the evidence against the former and lack of utility of the latter. There is debate within the scientific community about the relevance and importance of the hardening hypothesis particularly as the majority of smokers who have quit successfully since the 1960s, have done so without any formal support, including heavy smokers.^2^ The current balance of evidence is against the occurrence of hardening, and a useful way forward is perhaps to consider that, given the scale and consistency of the evidence against it, it is unlikely that large amounts of supportive evidence will emerge in the near future.

Hard-core smokers comprised around 2% of the total Australian adult population in 2010, and 1.2% of Victorians aged 26 and older in 2016 and represent a minority of smokers -- an estimated 10.7% of adult Australian smokers in 2010, and 9.1% of adult Victorian smokers in 2016.^20 21^ There have been calls to abandon the ‘hard-core’ smoker concept, in part because it perpetuates the stigma of smokers who continue to smoke, but also because it is not necessarily helpful in recognising and addressing the complex factors related to ongoing smoking.^10 26^

Comprehensive and multifaceted tobacco control measures have proved effective in reducing the prevalence of smoking in many countries, including Australia. These measures include smoke-free policies, mass media campaigns, plain packaging, graphic health warnings on packaging, price increases, and prohibitions on tobacco advertising, promotion and sponsorship.^27 28^ Such measures have made it harder for smokers to continue smoking large numbers of cigarettes throughout the day by reducing opportunities to smoke, making it very expensive to do so, and reducing its social acceptability. Brought together, these measures increase smokers’ motivation to quit and reduce their opportunities to smoke.

In parallel with reductions in the general population, the prevalence of smoking has fallen in many groups with historically high prevalence, including Aboriginal and Torres Strait Islander peoples.^29^ Nevertheless, the prevalence of current smoking remains disproportionately high in important parts of the Australian population, including Aboriginal and Torres Strait Islander communities, socioeconomically disadvantaged groups, and people affected by serious mental illness. Where softening is occurring among the general population of smokers, it may be occurring to a varying extent within important subpopulations, such as smokers from low socioeconomic backgrounds and smokers experiencing psychological distress.^5^ While one study found that the odds of being a hard-core smoker in Australia declined over the study years to a greater extent among those from high compared to low socioeconomic status groups,^21^ another study did not find a consistent pattern that indicated hardening over time in Victoria in any particular subgroup when examining age, gender, socioeconomic status or education.^20^ In disadvantaged populations, higher smoking prevalence relates to a range of interacting psychological, social, economic and cultural factors.^30^ Irrespective of how smoking is characterised, tobacco control interventions should be equitable and aim to reduce smoking across all population groups, including the most disadvantaged.

This review focused on peer-reviewed published evidence designed specifically to address questions regarding the hardening hypothesis in the population of smokers. Only repeat large population-based cross-sectional studies from Australia and similar high-income countries with a gap of at least five years between data points were included, with multiple authors independently extracting data. For certain hardening indicators, such as cigarettes per day and quit ratios, there are likely to be additional data in other publications, including government and technical reports. However, such publications often do not test statistically for change in indicators of hardening and may not be peer-reviewed.

The majority of primary research studies included in this review were of good quality. Most studies adjusted for relevant potential confounding factors over time, namely age and sex. Two studies adjusted for concomitant nicotine administration in the form of snus and nicotine replacement therapy^23 25^ which is relevant for the measurement of cigarettes per day if the use of concomitant nicotine-containing products has changed over time. A common limitation when assessing study quality was determining the validity of measures being used to assess hardening. Heterogeneity in the definitions and measurement of hardening indicators across studies makes it difficult to reliably ascertain the prevalence of hardened smoking in a population and to compare between studies and over time.^6 10 31^ This review examined patterns of hardening across a range of indicators in studies with data representing millions of people, and included high quality Australian national and state-level representative population-based survey data spanning two decades. Despite the variability in the definitions of hard-core smoker observed in the primary research, sensitivity analyses support the findings of a lack of hardening regardless of the definition of hard-core smoker used. Unlike other recent reviews of the hardening hypothesis,^6 9^ the authors of this review do not have any competing interests.

In conclusion, declining smoking prevalence in Australia and similar high-income countries has been accompanied by softening within the smoking population, characterised by increasing motivation to quit and reduced dependency; this is generally consistent with international evidence of softening or a lack of hardening over time. These findings indicate the effectiveness of ongoing tobacco control measures in reducing the prevalence of smoking as well as increasing motivation to quit and reducing dependency among the population that continues to smoke. Hence, based on the weight of the available evidence from high-income countries, the “hardening hypothesis” should be rejected.

## Data Availability

Not applicable

